# Profiling Clinical Researchers Effectively using Embeddings and Clustering

**DOI:** 10.1101/2024.01.16.24300807

**Authors:** Anika Sharma, Maxim Edelson, Tsung-Ting Kuo

## Abstract

Effective researcher profiling is key to support rapid research team formation. We developed a profiling method using (1) widely accessible publication titles, (2) document embedding vector representations to consider background, and (3) both general and specific types of datasets. Our results showed that the most similar researchers have cosine similarities of 0.287/0.258. Our preliminary results can support biomedical informaticians to expedite collaborative clinical studies, enhance research quality, and eventually improve patient healthcare outcomes.

## Introduction

Connecting with potential collaborators is vital while conducting scientific research. Rapid team formation is key to obtaining successful results for world-wide research projects, interdisciplinary collaborations, and urgent situation research task forces. Thus, it is vital to come up with an efficient solution for connecting researchers that allows researchers to invest their time in conducting research rather than building teams. A core requirement to build an efficient team is to accurately profile researchers according to their backgrounds. However, profiling experts is challenging because of the various research disciplines and large number of researchers world-wide. There are a few facets yet to be considered by existing works: (A) Requiring broad types of information; complete researcher background information^1^ may not be widely accessible. (B) Relying on enumerated skills/characteristics (e.g., “keywords” in publications)^1^; keywords may falsely indicate similarity because they do not necessarily account for the complete context/meaning of the study. (C) Focusing on a single type of dataset; this assumption may overlook a researcher’s different strengths, for example, general field knowledge vs. topic expertise.

## Method

In this preliminary study, we utilize widely-accessible publication title information, embedding similarity computation methods, and both general and specific types of datasets to streamline research team formation. We first filtered paper titles from the PubMed Central (PMC)^**3**^ and COVID-19 Open Research Dataset (CORD-19)^2^ datasets. Then, we created a “document” (title in our study) embedding vector representation for each title and averaged the vectors per researcher per dataset. Next, for each researcher we used cosine similarity to measure how close each researcher’s work was to others dataset-wise. Finally, we performed hierarchical clustering to group the researchers based on their scores. To evaluate our system, we identified researchers among the experts listed in the Citations Statistics of Biomedical Informatics Researchers website^4^, using the following criteria: (1) published in PMC and in the CORD-19 dataset, and (2) contains at least five Google Scholar keywords, one being “precision medicine” (our example field).

## Results

We identified 414,698 PMC papers and 288,001 CORD-19 papers and extracted their titles, as well as 7 experts for our preliminary study. For PMC, WQW and EA have the highest similarity (0.287), and RC and DPK are the most dissimilar (-0.058). For CORD-19, RC and WQW are the most similar (0.258), and JD and DPK have the lowest similarity (-0.061). We found that highly similar researchers do not necessarily have overlapping keywords, particularly for the more general PMC dataset.

## Discussion

The max similarity values, 0.287/0.258, were positive and not too close to the ceiling of 1.0, which indicates that our model (accounting for researchers’ backgrounds) may work in practice, because no two researchers should be completely similar. In this preliminary work, we have yet to conduct a more in-depth evaluation performed on datasets with more researchers or multi-lingual publications, use other researcher differentiation methods, and to adopt qualitative metrics to validate our model and its results.

## Conclusions

We developed a prototype to profile researchers, streamline the team formation process, and facilitate international collaboration, interdisciplinary projects, and emergency situations. Based on these preliminary results, biomedical informaticians may consider incorporating this workflow based on embedding and clustering to expedite collaborative clinical studies, enhance research quality, and eventually improve patient healthcare outcomes.

## Data Availability

All data produced in the present work are contained in the manuscript

## Acknowledgements

The authors AS, ME, and T-TK were funded by the U.S. NIH (R00HG009680, R01EB031030, R01GM118609, R01HG011066, R01HL136835, RM1HG011558, T15LM011271, U24LM013755, and U54HG012510). The content is solely the responsibility of the author and does not necessarily represent the official views of the NIH. The funders had no role in study design, data collection and analysis, decision to publish, or preparation of the manuscript.

## References

1. Tseng T-LB, Huang C-C, Chu H-W, Gung RR. Novel approach to multi-functional project team formation. International Journal of Project Management. 2004;22(2):147–59.

2. Wang LL, Lo K, Chandrasekhar Y, et al. Cord-19: the covid-19 open research dataset. ArXiv. 2020.

3. PubMed central: National Library of Medicine; [cited 2023Mar5]. Available from: https://www.ncbi.nlm.nih.gov/.

4. McCoy A, Lin J. Citations statistics of biomedical informatics researchers [cited 2023Mar5]. Available from: https://allisonbmccoy.github.io/scholar-scraper/index-bmi.html.

